# Evaluating the Scale Functioning of The Health System Literacy Scale in Canada (HSL-CAN): A Partial Credit Model

**DOI:** 10.64898/2026.01.23.26344690

**Authors:** Anh Thu Vo, Ying Cao, Lixia Yang, Robin Urquhart, Yanqing Yi, Peizhong Peter Wang

## Abstract

**Background:** Adapted from the Navigational Health Literacy scale (HLS_19_ – NAV-HL), the 20-item Health System Literacy (HSL-CAN) was developed to measure health system literacy among Chinese Canadians. However, little is known about the functioning of its response categories.

**Method:** This study aimed to evaluate the proper functioning of the response categories using data from 681 adult Chinese Canadians aged 30 years or older who have resided in Canada for at least six months. Data were collected through a cross-sectional online survey. The partial credit model was utilized to evaluate the dimensionality of the scale, item- and category-level fit statistics, and the ordering of category thresholds.

**Results:** Findings supported the unidimensional construct of the scale. Most items met the item theory response (IRT) model requirements, demonstrating acceptable item fit statistics, except for three items with outfit-t statistics outside the acceptable range. Adjacent response category thresholds were increased monotonically, and threshold distances were within the recommended upper range, although relatively narrow distances were observed for several items. Person separation reliability and person separation index indicated good internal consistency and adequate discrimination among individuals with different level of health system literacy.

**Conclusion:** This study provides evidence supporting the unidimensional construct and a proper functioning of the 5-point Likert response scale, suggesting that the HSL-CAN is a psychometrically appropriate instrument for evaluating health system literacy among Chinese Canadians. Future studies are needed to examine the scale’s applications across more diverse populations in Canada.

## Introduction

The Canadian healthcare system is complex and fragmented, characterized by uncoordinated care among providers, siloed departments, and sophisticated regulatory structures [1,2]. Navigating such a healthcare system can be overwhelming and struggling for many Canadian patients and caregivers [3], particularly for immigrants, who often suffer additional difficulties such as language barrier, cultural differences, socioeconomic barriers, and limited health literacy [4]. Sufficient understanding of healthcare system and strong navigational skills can enable Canadian individuals to interact with the system more effectively [5].

To facilitate the assessment of health system literacy, the Health System Literacy-Canada (HSL-CAN) scale was developed by adapting Navigational Health Literacy (HLS_19_ – NAV-HL) instrument from the International Health Literacy Population Survey and was initially validated among Chinese Canadians [6,7]. While the original 12-item scale employed a 4-point Likert response format ranging from ‘very difficult’ to ‘very easy’ [6], the HSL-CAN comprises 20 items using a 5-point Likert scale [7]. The inclusion of a neutral response was intended to reduce the tendency to ‘force a choice’ between ‘easy’ or ‘difficult’ option, thereby accommodating respondents with genuinely neutral opinion on an item [8]. Neutral responses may also reflect a satisficing option among many respondents who lack adequate knowledge of or interest in the issue [9]. The ultimate purpose of the HSL-CAN is to serve as a reliable and valid measure of health system literacy among patients and caregivers in Canada. Hence a comprehensive evaluation of its psychometric properties is necessary.

Two main test theories are commonly utilized to evaluate the psychometric properties: Classical test theory and Item Response Theory [10]. The classical test theory is relatively simple, relies on fewer assumptions, and is suitable for small sample size; however, this approach is sample-dependent performance and does not model item-level characteristics such as item difficulty and person ability [10,11]. In contrast, Item response theory (IRT) extends beyond the classical test theory by validating the construct at both the scale and item level. IRT models estimates the probability of a respondent answering correct an item as a function of their latent trait (person ability) and items’ characteristics (e.g., item difficulty, discriminant) [12]. Information functions derived from IRT model, like Cronbach’s alpha coefficient in classical test theory, also describe how precisely an item or scale measure the underlying construct across different level of person trait [12].

Several common IRT models are used for polytomous responses, including graded response, generalized partial credit, partial credit, and rating scale [12,13]. In the graded response and generalized partial credit, item discrimination parameters are allowed to vary across items while they are constrained to be equal in the partial credit and rating scale model [12].

Additionally, the partial credit models allow different response categories from item to item, whereas the rating scale assumes that the category threshold across items is equal [14,15].

Although the HSL-CAN scale has demonstrated satisfactory measurement properties using the classical test theory [7], its functioning of the response scale has not yet been thoroughly examined. Concerns remain regarding whether the 5-point Likert response scale is appropriate as intended to differentiate individual across different levels of health system literacy. Given its flexibility and fewer restrictions on response category structure, this study used partial credit model to evaluate the appropriateness of scale response functioning and item properties of the HSL-CAN scale.

## Methodology

### Study population

This study used data from previous published validation study that applied classical test theory to evaluate the psychometric properties of the HSL-CAN scale [7]. The sample comprised 681 Chinese individuals aged 30 or older who have lived in Canada for at least six months. Data were collected through an online cross-sectional survey administered from March 11 to July 19, 2024, through WeChat and other social media platforms (e.g., Facebook groups of Chinese immigrants in Canada, Chinese Canadian community organization online platforms, emails). Participants were geographically distributed across Canada, with 50.07% aged 50-64 years and 55.07% identifying as women. Detailed sample characteristics was reported elsewhere [7].

### Data analysis

This study examined the essential IRT assumptions, including (a) unidimensionality, (b) comparison of nested models (partial credit model (PMC-1-PL) vs. generalized partial credit model – (GPCM-2-PL)), (c) local independence, and (d) monotonicity. Item fit statistics and IRT-based reliability indices, and the functioning of adjacent-categorical thresholds were examined.

### IRT assumptions

#### Unidimensionality

To apply partial credit model, the key assumptions of unidimensionality and local independence must be met [16]. The unidimensionality refers to the measurement of a single latent trait. Rasch residuals that are defined as the differences between observed responses and responses predicted by the partial credit model should obtain one construct under this assumption [16,17]. A principal component analysis of partial credit model residuals was used to check the single construct [15,17]. The unidimensionality assumption was considered satisfied when the first component extracted from the principal component analysis, known as first contrast, had an eigenvalue of < 2.0 because this value is interpreted as random noise rather than evidence of a second dimension [15,18].

#### Local independence

Local independence assumes that, conditional on the latent trait, responses to each item are statistically independent [16,19]. Violations occurs when residual correlations between items are significantly high, indicating item redundancy [19]. Local independence was evaluated using Yen’s Q3 value, which represents residual correlations between items [15,19]. Because absolute Q3 values are influenced by the number of items, we reported the mean absolute Q3 and relative Q3 [15,19]. The relative Q3 is defined as a difference between the maximum Q3 and the mean Q3 [15,19]. A value of 0.2 or above is considered as violation in local independence assumption [15,19].

#### Comparing nested models

A likelihood ratio test was used to examine whether adding discrimination factor (GPCM-2PL) significantly improves model fit compared to the more parsimonious model (PCM-1PL) [12]. The difference in -2*log-likelihood between the two nested models follows a Chi-square, with degrees of freedom equal to the difference in the number of estimated parameters per item between the two models [12]. A statistically significant test indicated that the more complex model made the model better fit [12].

#### Fit statistics for the partial credit model

Scale validity was evaluated using item fit statistics in the partial credit model. Item fit statistics included mean square fit statistics (infit-MNSQ, outfit-MNSQ), standardized fit statistics (infit-t, and outfit-t) [20]. Infit-MNSQ is a weighted residual statistic sensitive to unexpected responses near an individual’s ability level [14, 20, 21], whereas outfit-MNSQ is unweighted residuals and more sensitive to extreme values (outliers) [16, 21]. Infit-MNSQ or outfit-MNSQ values between 0.5 and 1.5 logits indicate acceptable model fit [22].

Infit-t and outfit-t statistics are standardized fit statistics by converting MNSQ fit statistics into a normal t-distribution [21]. Values between -2 and 2 indicate a goodness-of-fit model [21,23]. However, these statistics are sensitive to sample size, with inflated Type I error in large sample size [23]. According to Winsteps guidelines, interpretation of MNSQ statistics were prioritized over standardized fit statistics [24].

### The partial credit model reliability

Person separation reliability, conceptually analogous to Cronbach’s alpha in classical test theory, was used to evaluate the scale’s ability to distinguish between individuals on the latent trait [25]. A value of ≥ 0.75 indicates a good reliability and > 0.9 indicates a very good reliability [25]. Additionally, the person separation index provides an estimate of the spread of person ability on the latent trait continuum, with values of ≥ 1.5 indicating acceptable separation [25].

### The responding scale investigation

Threshold calibrations and category fit statistics were examined to detect potential disordered response categories [21]. A threshold represents a point on the latent trait at which the probability of choosing two adjacent responses is equal [21]. Thresholds are considered disordered when they fail to increase monotonically across response categories [21]. Distances between adjacent thresholds should be at least 1.0 logits and no more than 5 logits to ensure meaningful differentiation between categories and to prevent measure imprecision, respectively [14].

Category fit statistics were assessed using with infit-MNSQ or outfit-MNSQ values greater than 2.0 or less than -2.0, indicating excessive noise or misinformative response categories [14, 21]. Moreover, a Wright map, known as a Rasch person-item map was used to identify potential disordered thresholds.

Data were analysed using R, with several packages used, including the TAM package for examining unidimensionality, local dependence issues, category fit statistics, the functioning of the responding scale, and virtualize Wright Map [26]; the eRm package for examining the model of fit and model reliability [27]; and the mirt package for comparing two nested models [28].

### Ethical approval and consent to participant

This study obtained the approvals from the Interdisciplinary Committee on Ethics in Human Research (ICEHR) at Memorial University of Newfoundland (ICEHR No. 20241177-ME and ICEHR No. 20250592-ME).

All participants were requested to provide their consent through the online informed consent forms. The online inform consent form was designed following the TCPS2 guidelines.

Participation was voluntary and anonymous, and they had the right to withdraw from the study without any penalties. Identifying variables such as IP address, WeChat ID, or email were not included in the data analysis.

## Results

### IRT assumptions

#### Unidimensionality

The results of the principal component analysis of Rasch residuals supported the unidimensionality assumption. The eigenvalues of the first (eigenvalue=1.38) and second (eigenvalue=1.34) residual contrasts were below the cutoff of 2.0, indicating that the residual variances were random noises rather than a meaningful secondary dimension. The scree plot of residual PCA did not exhibit a significant elbow, and the first and second residual contrasts accounted for 6.9% and 6.7% of total residual variance (**Fig 1**). These findings suggest an absence of a considerable secondary dimension.

**Fig 1.**
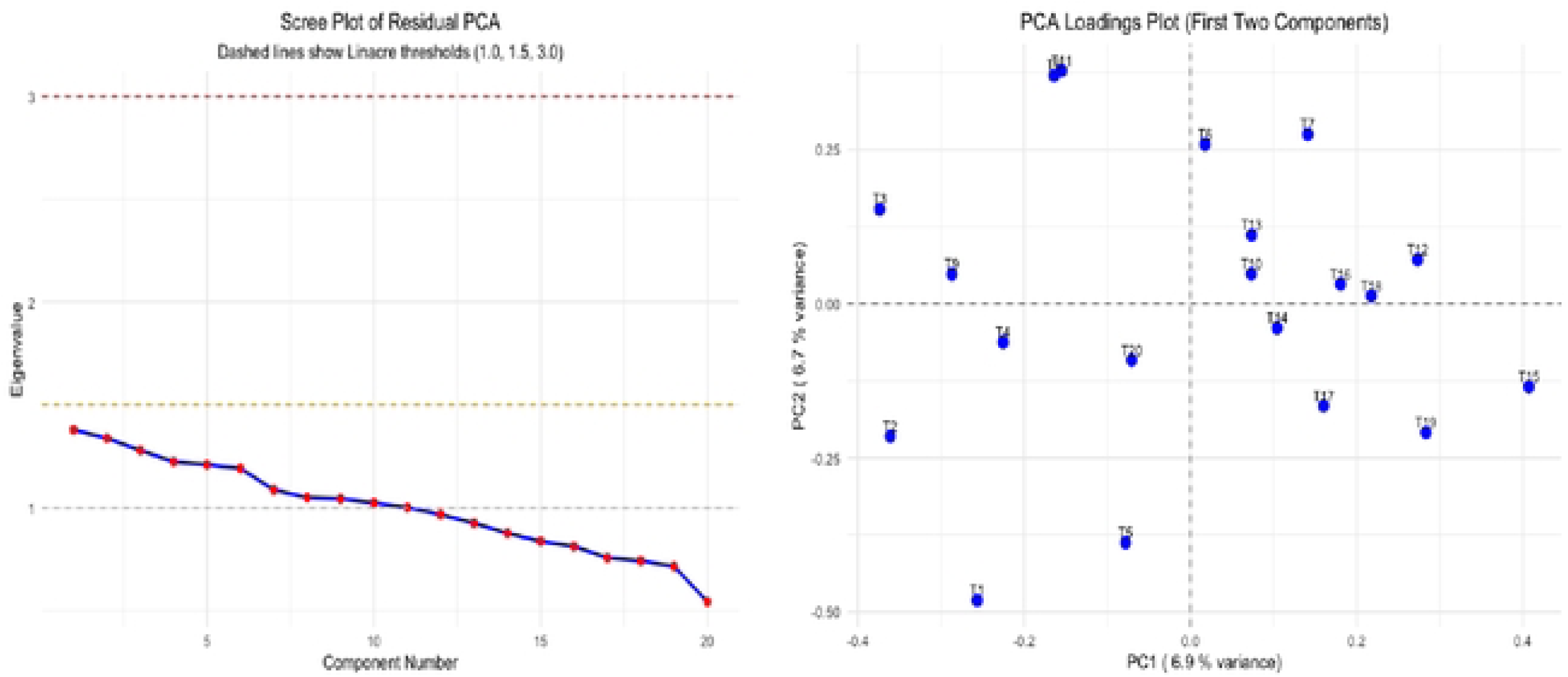
Scree Plot of Residual PCA and PCA Loading Plot for first two components of Rasch residuals

#### Comparing two nested models

The results of the likelihood ratio test are presented in **Table 1**. The test is not statistically significant difference in -2*log-likelihood between two nested models, indicating that the more parsimonious model (PCM-1PL) provided better model fit than the more complex model (GPCM-2PL) (-2*LL = *X*^2^= 14.967, df=19, p-value = 0.725).

**Table 1.** The likelihood ratio test to compare two nested models.

| Model | AIC | BIC | Log-likelihood | $\chi^2$ | df | p-value |
| --- | --- | --- | --- | --- | --- | --- |
| PCM-1PL | 40044.62 | 40411.02 | -19941.31 |  |  |  |
| GPCM-2PL | 40067.65 | 40520.00 | -19933.82 | 14.967 | 19 | 0.725 |

#### Local independence

Regarding the assumption of local independent, the absolute value of mean Q3 mean and the relative Q3 value were -0.045 and -0.131 (difference between mean = -0.045 and maximum value = 0.086). Both values were below the recommended benchmark of 0.2, indicating that the assumption of local independence was satisfied.

#### Item fit statistics of the partial credit model

**Table 2** presents the item difficulty estimates and fit statistics for all items in the HSL-CAN scale. Under the partial credit model, item difficulty represents the location of each item on the latent trait [13] and is calculated as the average of four category thresholds within an item [15]. Among 20 items, item 2 “*How easy is it for you to understand what different healthcare providers do?*” is considered as relatively easier (item location = -0.018 logit < 0) while the remaining items demonstrated average difficulty, with their locations around 0 logits [29].

**Table 2.**
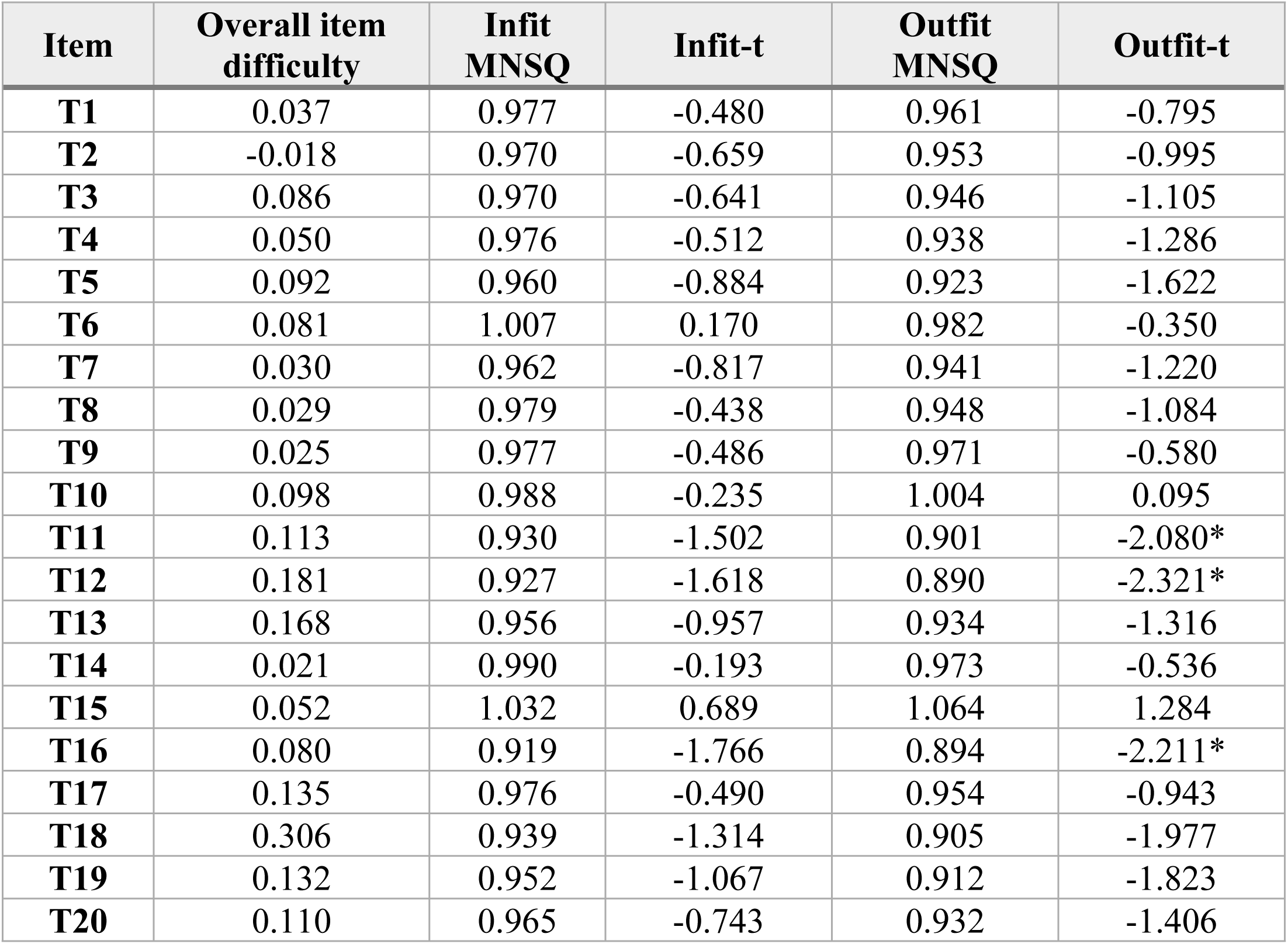
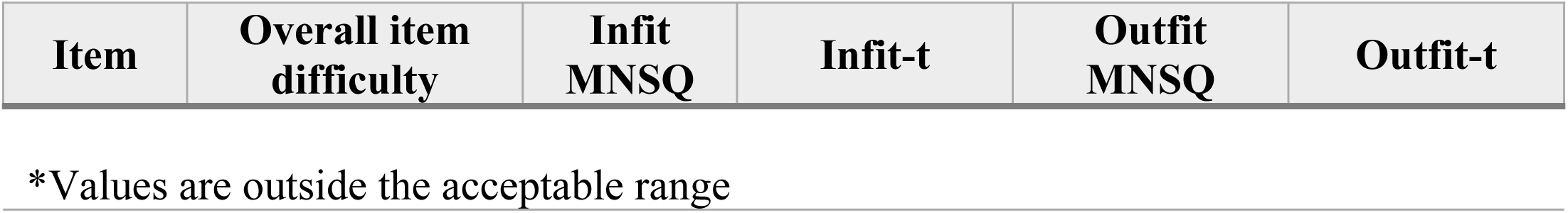
Item fit indices for the HSL-CAN scale.

| Item | Overall item difficulty | Infit MNSQ | Infit-t | Outfit MNSQ | Outfit-t |
| --- | --- | --- | --- | --- | --- |
| T1 | 0.037 | 0.977 | -0.480 | 0.961 | -0.795 |
| T2 | -0.018 | 0.970 | -0.659 | 0.953 | -0.995 |
| T3 | 0.086 | 0.970 | -0.641 | 0.946 | -1.105 |
| T4 | 0.050 | 0.976 | -0.512 | 0.938 | -1.286 |
| T5 | 0.092 | 0.960 | -0.884 | 0.923 | -1.622 |
| T6 | 0.081 | 1.007 | 0.170 | 0.982 | -0.350 |
| T7 | 0.030 | 0.962 | -0.817 | 0.941 | -1.220 |
| T8 | 0.029 | 0.979 | -0.438 | 0.948 | -1.084 |
| T9 | 0.025 | 0.977 | -0.486 | 0.971 | -0.580 |
| T10 | 0.098 | 0.988 | -0.235 | 1.004 | 0.095 |
| T11 | 0.113 | 0.930 | -1.502 | 0.901 | -2.080* |
| T12 | 0.181 | 0.927 | -1.618 | 0.890 | -2.321* |
| T13 | 0.168 | 0.956 | -0.957 | 0.934 | -1.316 |
| T14 | 0.021 | 0.990 | -0.193 | 0.973 | -0.536 |
| T15 | 0.052 | 1.032 | 0.689 | 1.064 | 1.284 |
| T16 | 0.080 | 0.919 | -1.766 | 0.894 | -2.211* |
| T17 | 0.135 | 0.976 | -0.490 | 0.954 | -0.943 |
| T18 | 0.306 | 0.939 | -1.314 | 0.905 | -1.977 |
| T19 | 0.132 | 0.952 | -1.067 | 0.912 | -1.823 |
| T20 | 0.110 | 0.965 | -0.743 | 0.932 | -1.406 |

\*Values are outside the acceptable range

All items obtained Infit MNSQ and outfit MNSQ values within the recommended ranges of 0.7 to 1.3 logit. Infit-t statistics for all items fell within an acceptable range of -2 to 2. While many outfit-t values were within this range, item 11 “*How easy is it for you to find support options that help you navigate the healthcare system?*”, item 12 “*How easy is it for you to decide a particular health service?*”, and item 16 “*How easy is it for you to find information on digital health services provided in your province?*” demonstrated outfit-t statistics below -2, indicating a potential overfit caused by extreme responses. However, these standardize fit statistics are sensitive to a large sample size, so considering that infit-MNSQ, outfit-MNSQ values, and infit-t statistics for all items within acceptable ranges, the partial credit model demonstrated an acceptable model fit.

### Reliability

Person separation reliability and person separation index are 0.855 and 2.426, respectively. The person separation reliability is greater than 0.75 and the separation index is greater than 1.5, indicating that the scale can differentiate individuals on the latent trait continuum.

### The responding scale investigation

Category thresholds and fit statistics of categories are demonstrated in **Table 3**. Thresholds increased monotonically across the scale, indicating that the category thresholds were not disordered (**S1 Fig**). Thresholds between categories 1→ 2, 2 → 3, 3 → 4, 4 → 5 ranged from - 1.610 to -1.050, -0.402 to -0.026, 0.387 to 0.552, respectively.

**Table 3.**
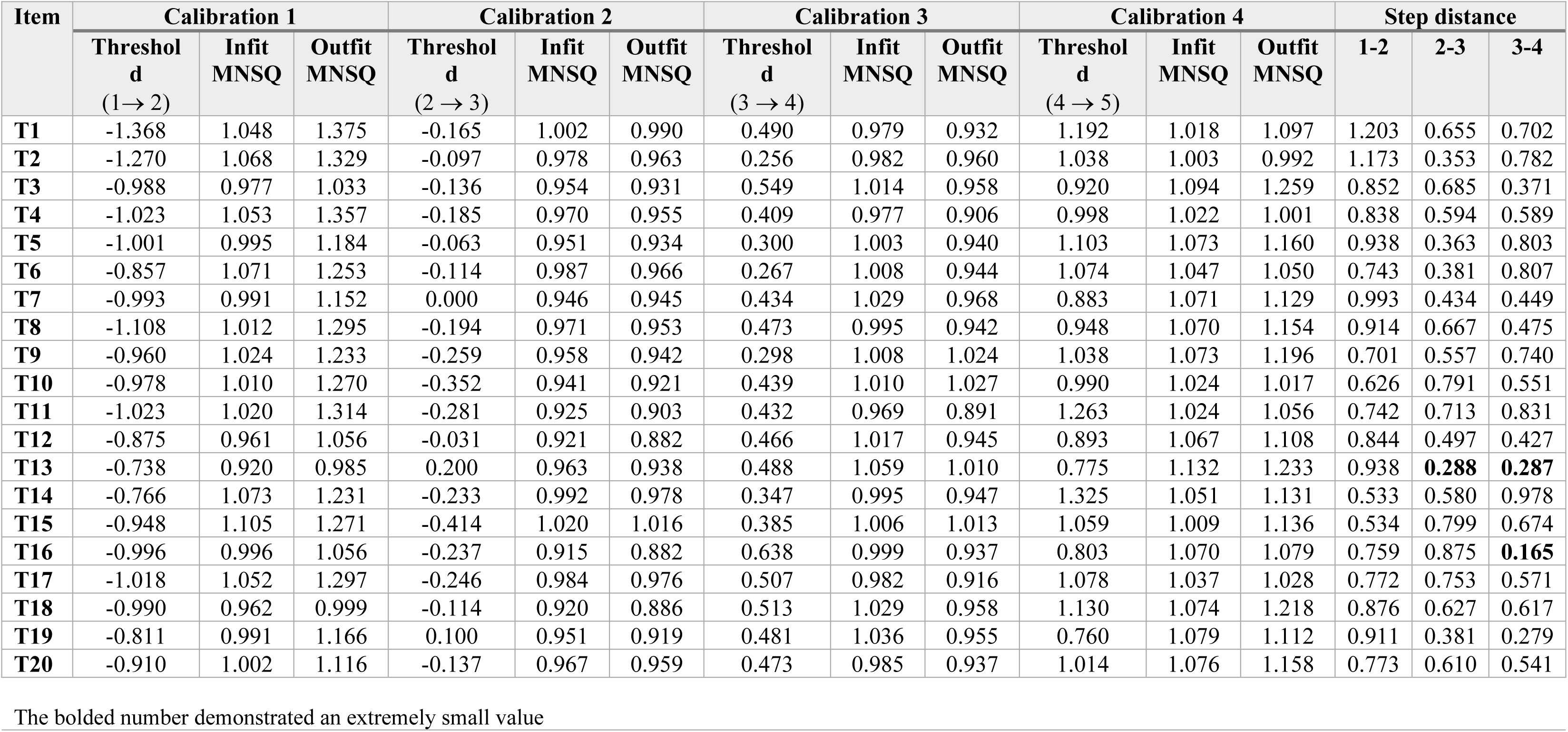
Category fit indices for the HSL-CAN scale.

The distances between the first and second thresholds ranged from 0.897 to 1.303 logits, with values below 1 observed for items D6, D9, D10, D14, and D15. Distances between the second and third threshold was below 1, ranging from 0.572 to 0.836 logits. Distances between the third and fourth threshold was from 0.724 to 1.131, with values below 1 observed for item 2-10, 12-13, 16-20.

**Fig 2** illustrates the category response curves for 5-point Likert scale, showing Item 1 with acceptable adjacent thresholds and item 13 with a narrow relatively adjacent threshold. All distances between adjacent thresholds were below 5.0 logit. Regarding category fit statistics, infit MNSQ and outfit MNSQ for all category thresholds fell within the acceptable ranges of 0.5 to 1.5.

**Fig 2.**
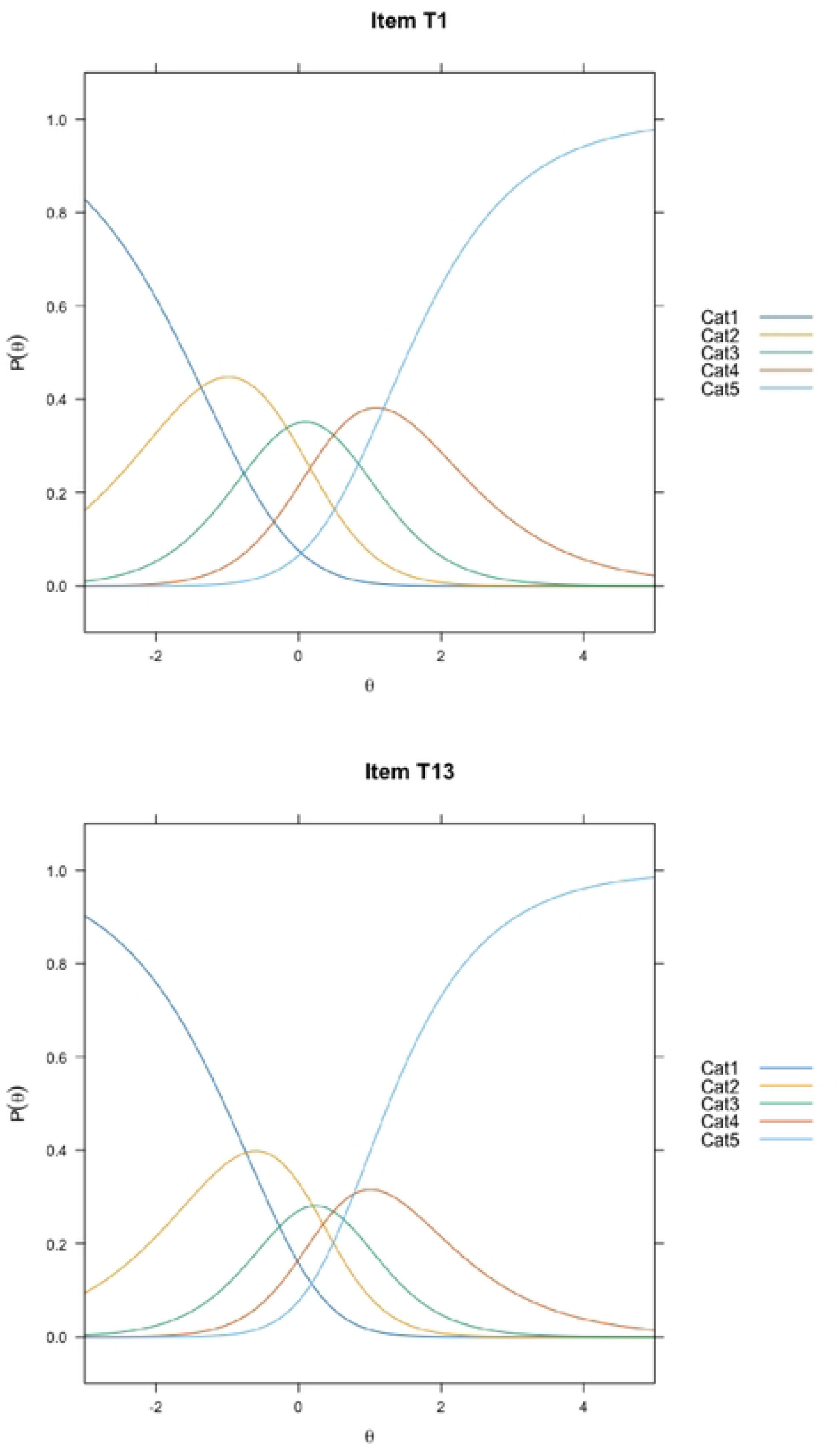
Category response curves for 5-point Likert HSL-CAN scale, Item 1 and item 13

Wright map illustrates the distribution of person abilities in the left-hand side and category thresholds on the right-hand side (**Fig 3**). Person abilities ranged from -3 to 2 logits, with most respondents clustered from -1 to 1 logits. Adjacent threshold for each item, labeled as Cat 1, Cat 2, Cat 3, Cat 4, were properly ordered, supporting the appropriate functioning of the response categories. No wide distances were observed between adjacent thresholds, although narrow distances were found in a few items (e.g., item 13). The category thresholds across items covered the range of person abilities, particularly in the moderate to high ability location (Cat 2 and Cat3), indicating that the scale effectively distinguish different person abilities with moderate to high ability levels (**Fig 3**).

**Fig 3.**
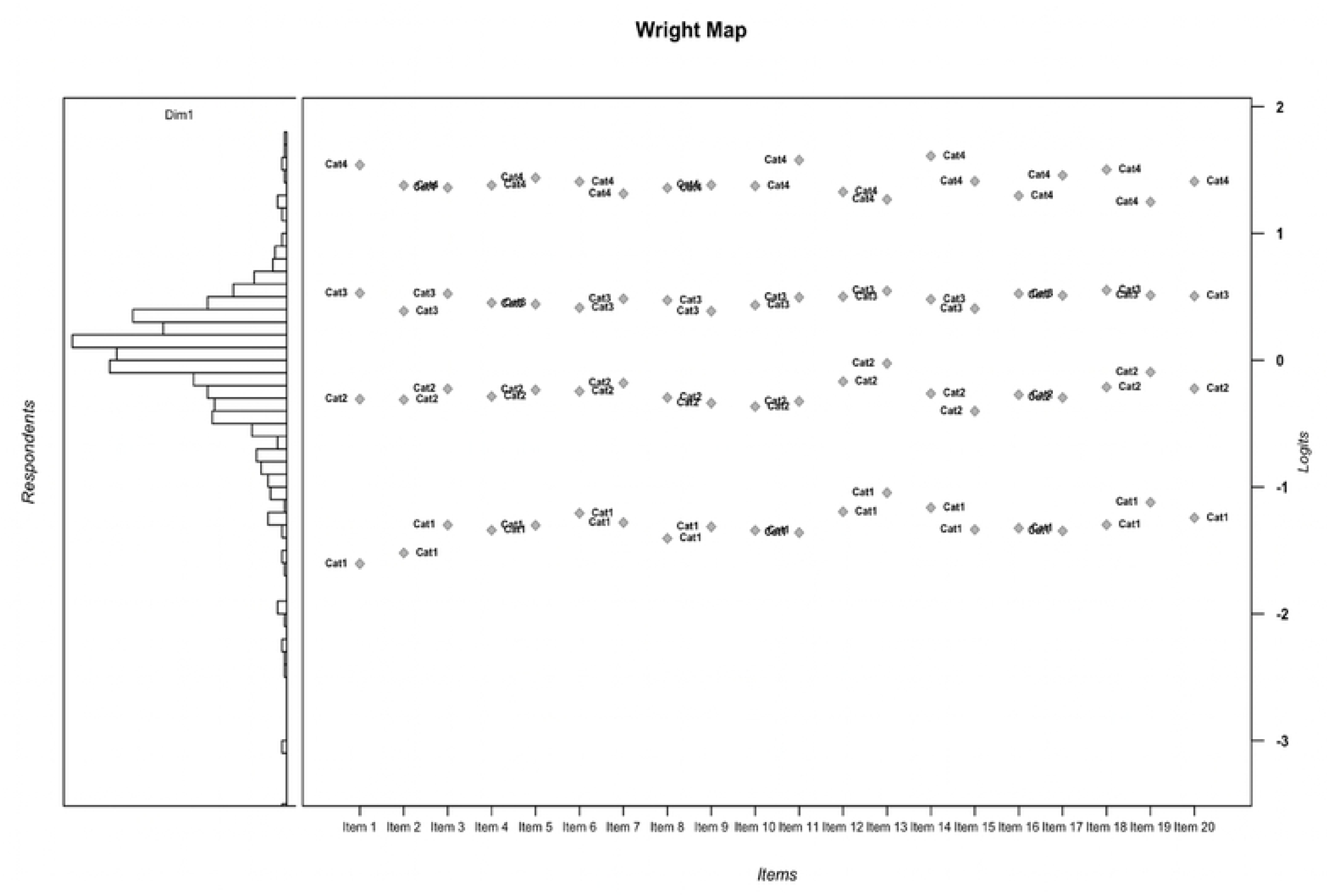
Wright Map

Overall, IRT assumptions as well as item-and category-level fit statistics were satisfied. Although the distances between adjacent thresholds for many items were below 1 logit, all distances were below 5.0 logits. Therefore, the 5-point Likert response scale is appropriate functioning.

### Revising category response analysis

Although category thresholds were appropriately ordered, a small step distance was identified between threshold 2 and 3 (corresponding to adjacent response categories 2-3 and 3-4) for item 13, and between threshold 3 and 4 (corresponding to adjacent response categories 3-4 and 4-5) for item 13 and 16. Given that the distance should be at least 1.0 logit between adjacent thresholds to obtain the proposed number of category responses [14], category responses were revised by collapsing categories. Three scenarios were examined: (A) collapsing category 3 (“neither easy nor difficult”) into category 2 (“somewhat difficult”), (B) collapsing category 3 into category 4 (“somewhat easy”), and (C) category 4 into category 5 (“extremely easy”). For each scenario, the ordering of category thresholds (**Fig S2, Fig S3, Fig S4**), item- and category-level fit indices (**S1 Table, S2 Table, S3 Table**), person separation reliability and person separation index were examined to determine whether any collapsing categories improved distance between thresholds compared to the original 5-point Likert response scale.

**Table 4** presents category thresholds and step distances between adjacent threshold for each scenario. Category thresholds were disordered in scenario A and C but remained ordered in scenario B. Therefore, the scenario A and C were excluded for further analysis. In scenario B, however, extremely narrow distances were observed between threshold 1 and 2 for item 14 (distance = 0.075) and 15 (distance = 0.105), indicating that the 4-point Likert response scale was not optimal.

**Table 4.** Category thresholds across three revised scale scenarios.

| Item | Scenario A<br>4-Likert response scale<br>(collapsing category 2 and category 3) |  |  |  |  | Scenario B<br>4-Likert response scale<br>(collapsing category 3 and category 4) |  |  |  |  | Scenario C<br>4-Likert response scale<br>(collapsing category 4 and category 5) |  |  |  |  |
| --- | --- | --- | --- | --- | --- | --- | --- | --- | --- | --- | --- | --- | --- | --- | --- |
|  | Threshold |  |  | Step distance |  | Threshold |  |  | Step distance |  | Threshold |  |  | Step distance |  |
|  | I | II | III | I→II | II→III | I | II | III | I→II | II→III | I | II | III | I→II | II→III |
| T1 | -2.033 | 1.144 | 1.279 | 3.177 | 0.135 | -1.418 | -0.600 | 2.136 | 0.818 | 2.736 | -1.434 | -0.162 | 0.239 | 1.272 | 0.401 |
| T2 | -1.902 | 0.944 | 1.123 | 2.846 | 0.179 | -1.316 | -0.624 | 1.862 | 0.692 | 2.486 | -1.335 | -0.094 | -0.037 | 1.241 | 0.057 |
| T3 | -1.647 | 1.212 | 1.006 | 2.859 | -0.206* | -1.027 | -0.546 | 1.900 | 0.481 | 2.446 | -1.044 | -0.126 | 0.220 | 0.918 | 0.346 |
| T4 | -1.701 | 1.053 | 1.085 | 2.754 | 0.032 | -1.062 | -0.649 | 1.901 | 0.413 | 2.550 | -1.081 | -0.178 | 0.103 | 0.903 | 0.281 |
| T5 | -1.623 | 0.999 | 1.188 | 2.622 | 0.189 | -1.036 | -0.572 | 1.946 | 0.464 | 2.518 | -1.057 | -0.056 | 0.025 | 1.001 | 0.081 |
| T6 | -1.504 | 0.943 | 1.160 | 2.447 | 0.217 | -0.891 | -0.636 | 1.902 | 0.255 | 2.538 | -0.913 | -0.107 | -0.015 | 0.806 | 0.092 |
| T7 | -1.590 | 1.161 | 0.966 | 2.751 | -0.195* | -1.029 | -0.454 | 1.798 | 0.575 | 2.252 | -1.047 | 0.011 | 0.093 | 1.058 | 0.082 |
| T8 | -1.790 | 1.113 | 1.035 | 2.903 | -0.078* | -1.151 | -0.633 | 1.887 | 0.518 | 2.520 | -1.167 | -0.187 | 0.152 | 0.980 | 0.339 |
| T9 | -1.673 | 0.912 | 1.126 | 2.585 | 0.214 | -1.000 | -0.767 | 1.885 | 0.233 | 2.652 | -1.021 | -0.255 | 0.005 | 0.766 | 0.260 |
| T10 | -1.739 | 1.014 | 1.080 | 2.753 | 0.066 | -1.021 | -0.804 | 1.913 | 0.217 | 2.717 | -1.039 | -0.348 | 0.131 | 0.691 | 0.479 |
| T11 | -1.749 | 1.034 | 1.353 | 2.783 | 0.319 | -1.065 | -0.736 | 2.176 | 0.329 | 2.912 | -1.085 | -0.277 | 0.199 | 0.808 | 0.476 |
| T12 | -1.488 | 1.177 | 0.977 | 2.665 | -0.200* | -0.908 | -0.473 | 1.825 | 0.435 | 2.298 | -0.927 | -0.019 | 0.128 | 0.908 | 0.147 |
| T13 | -1.259 | 1.309 | 0.855 | 2.568 | -0.454* | -0.762 | -0.233 | 1.716 | 0.529 | 1.949 | -0.783 | 0.218 | 0.111 | 1.001 | -0.107 |
| T14 | -1.473 | 0.967 | 1.414 | 2.440 | 0.447 | -0.798 | -0.723 | 2.190 | **0.075** | 2.913 | -0.822 | -0.227 | 0.130 | 0.595 | 0.357 |
| T15 | -1.739 | 0.935 | 1.149 | 2.674 | 0.214 | -0.991 | -0.886 | 1.953 | **0.105** | 2.839 | -1.011 | -0.412 | 0.097 | 0.599 | 0.509 |
| T16 | -1.703 | 1.256 | 0.890 | 2.959 | -0.366* | -1.038 | -0.616 | 1.838 | 0.422 | 2.454 | -1.053 | -0.228 | 0.269 | 0.825 | 0.497 |
| T17 | -1.728 | 1.123 | 1.166 | 2.851 | 0.043 | -1.061 | -0.673 | 2.033 | 0.388 | 2.706 | -1.078 | -0.240 | 0.225 | 0.838 | 0.465 |
| T18 | -1.641 | 1.184 | 1.217 | 2.825 | 0.033 | -1.027 | -0.540 | 2.085 | 0.487 | 2.625 | -1.046 | -0.105 | 0.246 | 0.941 | 0.351 |
| T19 | -1.371 | 1.252 | 0.842 | 2.623 | -0.410* | -0.838 | -0.338 | 1.700 | 0.500 | 2.038 | -0.858 | 0.113 | 0.098 | 0.971 | -0.015 |
| T20 | -1.571 | 1.135 | 1.102 | 2.706 | -0.033* | -0.945 | -0.579 | 1.949 | 0.366 | 2.528 | -0.964 | -0.128 | 0.172 | 0.836 | 0.300 |
| Conclusion | Disordered |  |  |  |  | Ordered |  |  |  |  | Disordered |  |  |  |  |

Category-level fit indices were satisfied across all response category and items, indicating the category functioning was generally appropriate. However, at the item level, item 18 (“*How easy is it for you to find out if your health insurance covers visits to certain health providers*”) demonstrated outfit-t statistics below -2 while item 12 “*How easy is it for you to decide a particular health service?*” exhibited infit-t and outfit-t statistics outside acceptable ranges, suggesting misfit (see Supplementary S3 – Table S3.1). Moreover, person separation reliability (0.812) and person separation index (2.076) in scenario B were smaller than those observed for the original 5-point Likert response scales (0.855 and 2.426, respectively), suggesting that the collapsing category 3 into category 4 reduced the scale’s ability to differentiate between respondents with different level of health system literacy. In conclusion, the 5-point Likert scale illustrated the most appropriate functioning in this study, based on ordered category thresholds, acceptable category- and item-level fit indices, and good reliability indexes.

## Discussion

This study provides evidence supporting the appropriateness of the 5-point scale for the HSL-CAN adapted from the HLS_19_ – NAV-HL, using the partial credit model. By using the partial credit model, the study could examine item-level psychometric properties, including person ability, item difficulty, and the functioning of scale responses along the latent trait continuum.

The unidimensionality of the scale was confirmed by the principal component analysis of Rasch residuals. The first contrast eigenvalue (1.38) was below the accepted threshold of 2.0 logits and accounted for only 6.9% of the total variance, suggesting that the residual variance did not represent a meaningful secondary dimension. The findings reinforce the unidimensional structure reported previously published study [7]. Our findings are also consistent with those of Touzani et al. (2024), who demonstrated a unidimensional construct for the original HLS19-NAV scale using the partial credit model, reporting the first residual contrast’s eigenvalue of 1.188 that explained 15% of the total variance [30]. In contrast, findings in Griese et al. (2022) weakly supported the unidimensionality because the statistically significant number of t-test for person ability estimated from system level subscale and that from organizational level subscale were greater than 5% across European countries, except for from Czech Republic [6].

Differences in the scale dimensionality across studies may reflect variations in contexts in which these studies were conducted, as well as difference in language and population characteristics. Although IRT aims to estimate relatively sample-invariant item parameters once model fit is established [12], measurement properties can vary across different samples populations. Therefore, re-evaluation of psychometric properties can be recommended when the scale is applied to new populations with different cultural background, even when IRT is employed.

The original HSL_19_-NAV uses a 4-point Likert scale (1-very difficult, 2-difficult, 3-easy, 4-very easy) [6, 30, 31]. In contrast, the HSL-CAN exploited a 5-point Likert scale ranging from “1-Extremely difficult” to “5-Extremely easy”, incorporating a neutral midpoint [7]. According to IRT assumption, a properly functioning of 5-point Likert scale (1-“extremely difficult”, 2-“somewhat difficult”, 3- “neither easy nor difficult”, 4-“somewhat easy”, 5-“extremely easy”) must follow monotonically ordered category thresholds, indicating that respondents with higher ability are more likely to endorse higher response categories. When neutral responses do not reflect true ability, neutral bias may result in category thresholds disordered [9, 32]. In this study, the findings from the partial response model demonstrated that the category thresholds are properly ordered, supporting the appropriateness of the response scale.

Relatively narrow distances between adjacent thresholds were observed for several items. Narrow adjacent threshold distances can reduce the measurement precision when because the respondent abilities cluster closely across adjacent categories [13, 20]. Conversely, excessively broad adjacent threshold distances can limit the scale to distinguish different person ability level on the latent trait [13, 20]. In the present study, threshold distances across items fell within the acceptable upper threshold of 5 logits. Additionally, most items and category fit statistics fell within the recommended range of 0.5 to 1.5 logit. The findings suggest that the HSL-NAV with 5-point Likert scale demonstrated acceptably functioning response categories, even though threshold distances should be examined in future applications.

Regarding the scale reliability, the partial credit model yielded a person separation reliability of 0.855 and a person separation index of 2.426, indicating good internal consistency and adequate discrimination among participants with different ability level of health system literacy on the latent trait continuum. Similarly, high person separation index values were reported in Griese et al. (2022) and Touzani et al. (2024) studies [6, 30].

Taken together, careful investigation of category thresholds and fit indices ensure that the functioning of response categories is proper to meaningful discriminate person ability levels.

In practice, instrument selection often prioritizes several main psychometric properties, including content validity, structural validity, construct validity, and reliability, while the functioning of response categories received relatively less attention. Neglecting response scale evaluation can fail to accurately estimate person ability or to detect disordering issues arising from response bias, ultimately affecting the scale applications.

### Strengths and limitations

This study provides strong evidence supporting the appropriateness of the 5-point Likert response scale. The partial credit model allows to examine the psychometric properties at the item level, including item difficulty and person ability. This approach can address major limitations of the classical test theory that primarily evaluates measurement properties based on a total scale score.

Several limitations need to be acknowledged. First, data were collected using a convenient sampling procedure, so selection bias must be considered. Based on the person-item map, the HSL-CAN appears to be more informative for individuals with moderate to higher levels of health system literacy, indicating potential limitations in measurement precision at lower ability levels. However, nearly 98% of participants in this study were immigrants, with more than 75% having resided in Canada for at least 10 years. Most participants had relatively high educational levels, with over 50% getting a master’s or doctoral’s degree, which may reflect Canadian immigrant policy [33]. These characteristics suggest that the scale can be suitable tool for assessing health system literacy among established immigrants in Canada, but further research is required to evaluate its applicability among individuals with lower educational attainment or recent immigrants.

Second, the study sample included only participants aged 30 or older due to cultural considerations and survey requirements [7]. Consequently, little is known about the scale performance on younger populations. Future studies should examine psychometric properties of the scale across broader populations from diverse background to strengthen evidence for its generalizability.

## Conclusion

In summary, the proper functioning of 5-point Likert response scale of the HSL-CAN is proved from the partial credit model. All items satisfied main IRT assumptions and demonstrated adequate ability to distinguish individuals in the latent trait continuum. However, although the adjacent thresholds were properly ordered and item fit indices were within acceptable ranges, relatively narrow threshold distances were observed for several items, suggesting some limitations in category differentiation. Moreover, the scale is suitable for measuring health system literacy among people aged 30 or older with moderate to high ability level. Further validation studies are needed to examine the scale’s performance across broader populations in Canada.

## Acknowledgments

The authors would like to thank M-POHL Action Network and its affiliated projects for the approval granted to utilize the HLS19-NAV. We would like to express our gratitude for all participants and our collaborating organizations for promoting the project. We wish to express our sincere appreciation to Nan Lei whose support in verifying the Chinese survey versions.

## Authors’ contributions

- Conceive and conceptualization: ATV
- Data collection: ATV, YC, PW
- Scale translation: YC, ATV
- Data analysis, interpretation, drafting original manuscript: ATV
- Writing original draft: ATV, RU, LY, YY, YC, PW
- Project administration: YC
- Project supervision: LY, RU, YY, PW
- All authors read and approved the final manuscript.

## Data availability Statement

Data used in this study are available on the request from the corresponding author.

